# Humoral responses against BQ.1.1 elicited after breakthrough infection and SARS-CoV-2 mRNA vaccination

**DOI:** 10.1101/2022.12.20.22283723

**Authors:** Alexandra Tauzin, Mehdi Benlarbi, Halima Medjahed, Yves Grégoire, Josée Perreault, Gabrielle Gendron-Lepage, Laurie Gokool, Chantal Morisseau, Pascale Arlotto, Cécile Tremblay, Daniel E. Kaufmann, Valérie-Martel-Laferrière, Inès Levade, Marceline Côté, Gaston de Serres, Renée Bazin, Andrés Finzi

**Affiliations:** Centre de Recherche du CHUM, Montreal, QC, Canada; Département de Microbiologie, Infectiologie et Immunologie, Université de Montréal, Montreal, QC, Canada; Héma-Québec, Affaires Médicales et Innovation, Quebec, QC, Canada; Département de Médecine, Université de Montréal, Montreal, QC, Canada; Division of Infectious Diseases, Department of Medicine, University Hospital of Lausanne and University of Lausanne, Lausanne, Switzerland; Laboratoire de Santé Publique du Québec, Institut National de Santé Publique du Québec, Sainte-Anne de-Bellevue, QC, Canada; Department of Biochemistry, Microbiology and Immunology, and Centre for Infection, Immunity, and Inflammation, University of Ottawa, Ottawa, ON, Canada; Institut National de Santé Publique du Québec, Quebec, QC, Canada

**Keywords:** COVID-19, SARS-CoV-2, mRNA bivalent vaccine, Hybrid immunity, Humoral responses, BQ.1.1

## Abstract

The Omicron BQ.1.1 variant is now the major SARS-CoV-2 circulating strain in many countries. Because of the many mutations present in its Spike glycoprotein, this variant is resistant to humoral responses elicited by monovalent mRNA vaccines. With the goal to improve immune responses against Omicron subvariants, bivalent mRNA vaccines have recently been approved in several countries. In this study, we measure the capacity of plasma from vaccinated individuals, before and after a fourth dose of mono-or bivalent mRNA vaccine, to recognize and neutralize the ancestral (D614G) and the BQ.1.1 Spikes. Before and after the fourth dose, we observe a significantly better recognition and neutralization of the ancestral Spike. We also observe that fourth-dose vaccinated individuals who have been recently infected recognize and neutralize better the BQ.1.1 Spike, independently of the mRNA vaccine used, than donors who have never been infected or have an older infection. Our study supports that hybrid immunity, generated by vaccination and a recent infection, induces higher humoral responses than vaccination alone, independently of the mRNA vaccine used.

## Introduction

The Omicron BQ.1.1 variant is a sublineage of the BA.5 variant that spreads very rapidly and is now the major circulating lineage in several countries [1,2]. Recent studies have shown that original SARS-CoV-2 mRNA vaccines, based on the ancestral Wuhan strain Spike (S), lead to poor humoral responses against several Omicron subvariants, including the BQ.1.1 variant [3–5]. With the goal to improve immune responses against these subvariants, Moderna and Pfizer bivalent vaccines have recently been approved by health authorities in many countries [6–8]. These updated versions of the vaccines are composed of mRNA coding for the expression of both the ancestral and an Omicron subvariant S [9,10]. However, the continued evolution of SARS-CoV-2 has resulted in the emergence of multiple Omicron sub-lineages showing signs of convergent evolution by the acquisition of the same immune escape mutation in the RBD region of the Spike protein. Notably, all five recent convergent mutations are present in BQ. 1.1: R346T, K444T, L452R, N460K, or F486V [3]. Because of these newly acquired mutations, the benefits of bivalent compared to monovalent vaccines against this lineage remain to be established.

It is well accepted now that hybrid immunity leads to better immune responses and protection from severe outcomes than vaccination alone [11–17]. Because the original mRNA vaccines poorly prevent viral transmission, an important part of the vaccinated population have been recently infected by Omicron subvariants, leading to improved immune responses in these individuals compared to SARS-CoV-2 naïve individuals who have just been vaccinated.

In this study, we evaluated the capacity of plasma antibodies to recognize and neutralize the original D614G and the Omicron BQ.1.1 subvariant S four weeks (W4-Va3) and four months (M4-Va3) after the third dose and four weeks after the fourth dose (W4-Va4) of mRNA vaccines (Figure 1A). These participants mainly received as their first three doses of vaccine the Pfizer monovalent vaccine, and as the fourth dose either the Pfizer or Moderna monovalent or Pfizer (BA.4/5) or Moderna (BA.1) bivalent vaccines. We also measured the anti-nucleocapsid (N) level to determine if the donors had recent breakthrough infection (BTI), i.e., they have been infected between their third and fourth doses of vaccine by a Omicron sublineage. Basic demographic characteristics of the cohort are summarized in Table 1.

**Table 1.**
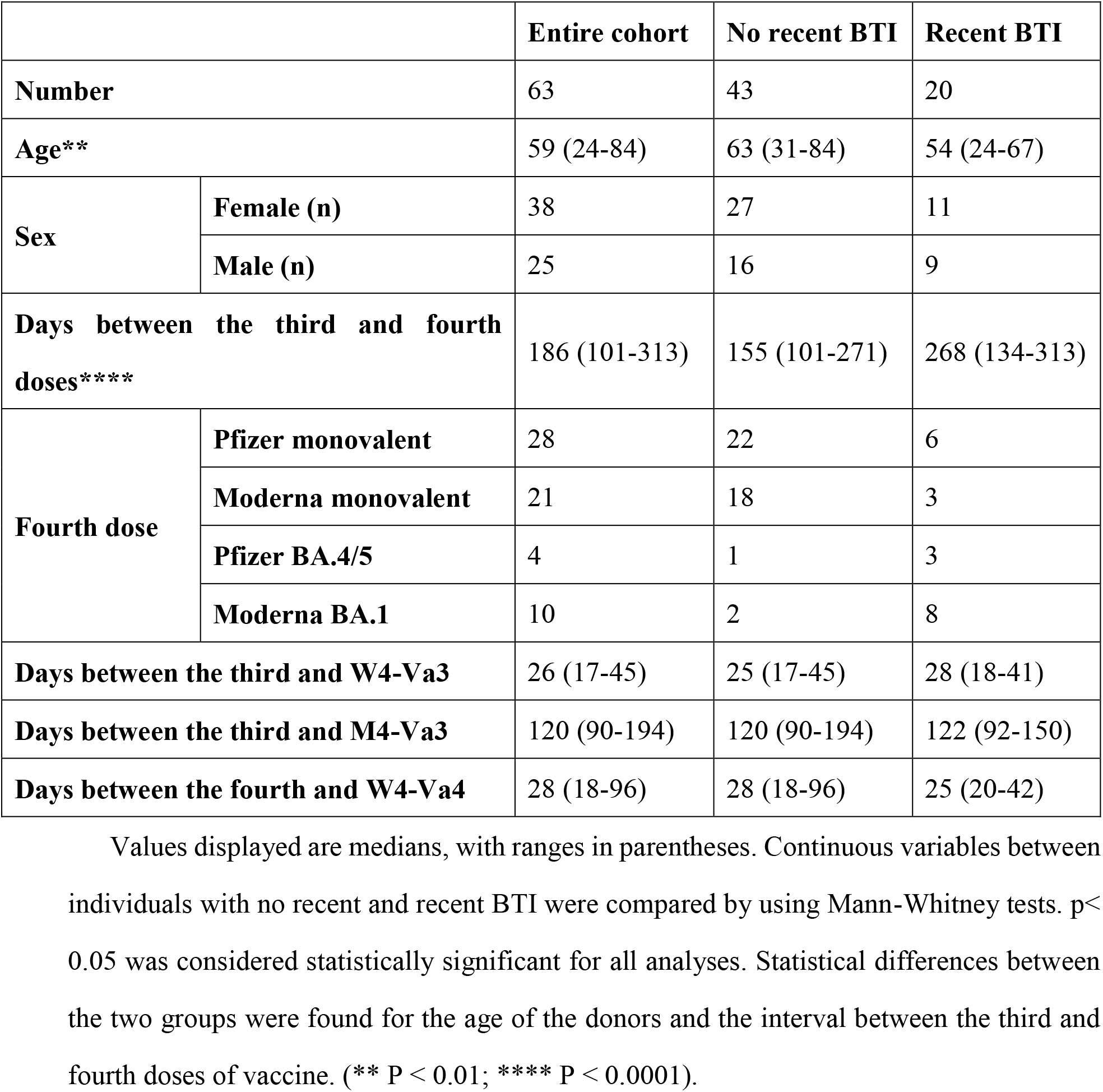
Characteristics of the SARS-CoV-2 vaccinated cohort.

|  |  | **Entire cohort** | **No recent BTI** | **Recent BTI** |
| --- | --- | --- | --- | --- |
| **Number** |  | 63 | 43 | 20 |
| **Age\*\*** |  | 59 (24-84) | 63 (31-84) | 54 (24-67) |
| **Sex** | **Female (n)** | 38 | 27 | 11 |
|  | **Male (n)** | 25 | 16 | 9 |
| **Days between the third and fourth doses\*\*\*\*** |  | 186 (101-313) | 155 (101-271) | 268 (134-313) |
| **Fourth dose** | **Pfizer monovalent** | 28 | 22 | 6 |
|  | **Moderna monovalent** | 21 | 18 | 3 |
|  | **Pfizer BA.4/5** | 4 | 1 | 3 |
|  | **Moderna BA.1** | 10 | 2 | 8 |
| **Days between the third and W4-Va3** |  | 26 (17-45) | 25 (17-45) | 28 (18-41) |
| **Days between the third and M4-Va3** |  | 120 (90-194) | 120 (90-194) | 122 (92-150) |
| **Days between the fourth and W4-Va4** |  | 28 (18-96) | 28 (18-96) | 25 (20-42) |

Values displayed are medians, with ranges in parentheses. Continuous variables between individuals with no recent and recent BTI were compared by using Mann-Whitney tests. $p <$ 0.05 was considered statistically significant for all analyses. Statistical differences between the two groups were found for the age of the donors and the interval between the third and fourth doses of vaccine. (\*\* $P < 0.01$ ; \*\*\*\* $P < 0.0001$ ).

**Figure 1.**
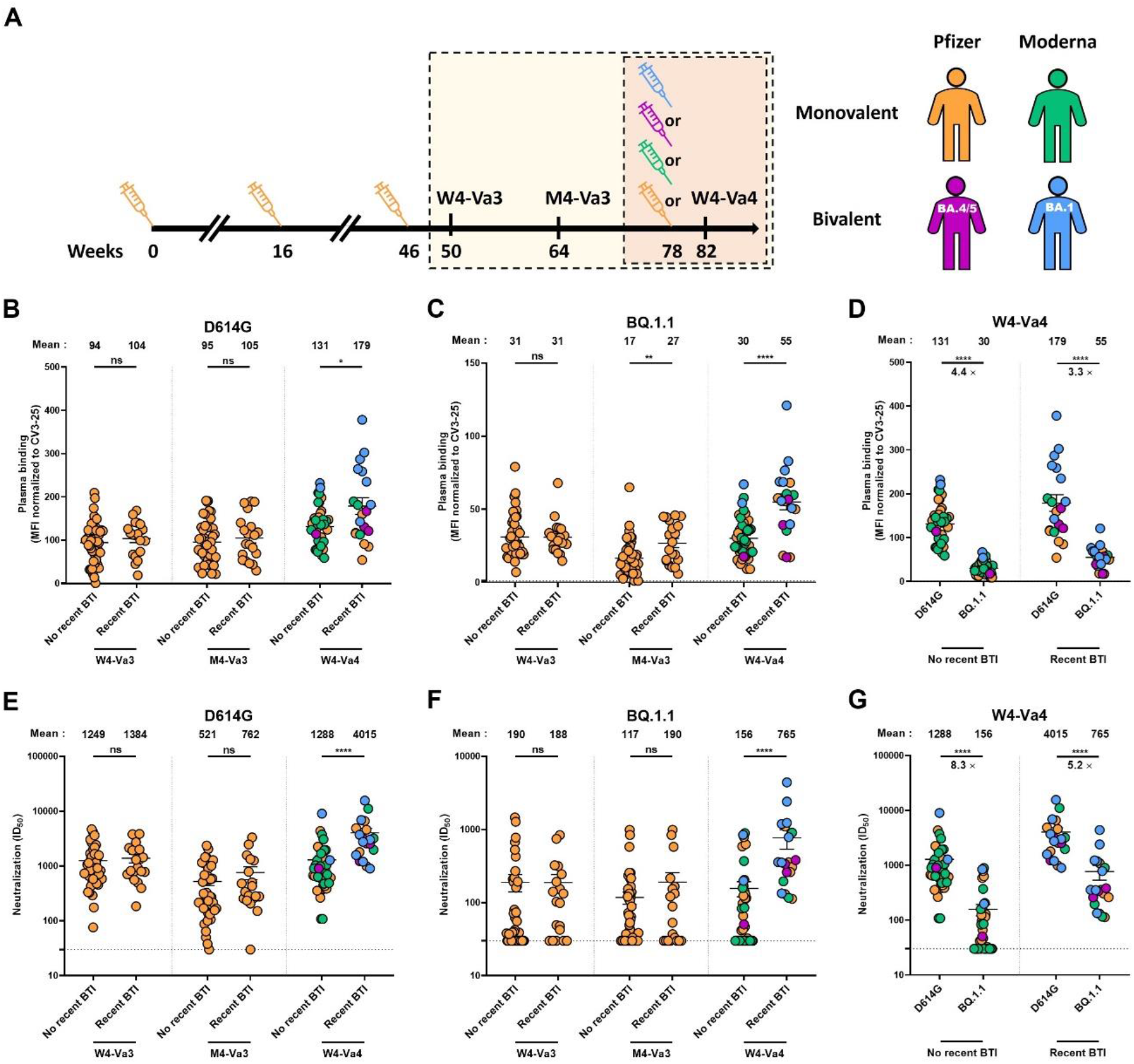
Recognition and neutralization of the D614G and BQ.1.1 Spikes after the third and fourth doses of SARS-CoV-2 vaccine in individuals with or without a recent breakthrough infection. (**A**) SARS-CoV-2 vaccine cohort design. The yellow box identifies the three timepoints under study shown in panels B, C, E and F and the red box the period presented in panels D and G. (**B-D**) 293T cells were transfected with the full-length D614G or BQ.1.1 S, stained with the CV3-25 mAb or with plasma from vaccinated individuals and analyzed by flow cytometry. The values represent the MFI normalized by CV3-25 mAb binding. (**E-G**) Neutralization activity was measured by incubating pseudoviruses bearing SARS-CoV-2 S glycoproteins, with serial dilutions of plasma for 1 h at 37°C before infecting 293T-ACE2 cells. Neutralization half maximal inhibitory serum dilution (ID_50_) values were determined using a normalized non-linear regression using GraphPad Prism software. Individuals vaccinated with Pfizer monovalent, Moderna monovalent, Pfizer bivalent (BA.4/5) or Moderna bivalent (BA.1) fourth dose are represented by orange, green, purple and blue points respectively. Limits of detection are plotted. Error bars indicate means ± SEM. (* P < 0.05; ** P < 0.01; **** P < 0.0001; ns, non-significant).

## Materials and Methods

### Ethics statement

The study was conducted in accordance with the Declaration of Helsinki in terms of informed consent and approval by an appropriate institutional board. The protocol was approved by the Ethics Committee of CHUM (19.381, approved on February 28, 2022) and Héma-Québec (2022-016, approved on October 7, 2022).

### Human subjects

The study was conducted in 63 individuals (25 males and 38 females; age range: 24-84 years). 20 of these individuals had recent breakthrough infection with an Omicron sublineage (9 males and 11 females; age range: 24-67 years), i.e. as determined by the increase in anti-N levels between W4-Va3 and M4-Va3 or between M4-Va3 and W4-Va4 using a recently described analytical approach [18] (Figure S1). For the other donors (16 males and 27 females; age range: 31-84 years), we did not observe a significant increase of the anti-N levels, although some of them have a history of infection. No other specific criteria such as number of patients (sample size), sex, clinical or demographic were used for inclusion.

### Plasma samples and antibodies

Plasma samples were either recovered from whole blood or directly obtained from the PlasCov biobank [19], heat-inactivated for 1 hour at 56°C and stored at -80°C until use in subsequent experiments. Pre-pandemic plasma samples were used as negative controls in cytometry assays (data not shown). The conformationally independent S2-specific monoclonal antibody CV3-25 was used as a positive control and to normalize Spike expression in flow cytometry assays, as described [4,20–23]. Alexa Fluor-647-conjugated goat anti-human antibodies (Abs) able to detect all Ig isotypes (anti-human IgM+IgG+IgA; Jackson ImmunoResearch Laboratories, Cat # 109-605-064) were used as secondary Abs to detect plasma binding in flow cytometry experiments.

### Plasmids

The plasmids encoding the SARS-CoV-2 D614G and BQ.1.1 Spike variants were previously described [4]. The pNL4.3 R-E-Luc plasmid was obtained from the NIH AIDS Reagent Program (Cat# 3418). The pIRES2-EGFP expressing plasmid was purchased from Clontech (Cat# 6029-1).

### Cell lines

293T human embryonic kidney cells (obtained from ATCC, Cat# CRL-3216) were maintained at 37°C under 5% CO_2_ in Dulbecco’s modified Eagle’s medium (DMEM) (Wisent) containing 5% fetal bovine serum (FBS) (VWR) and 100 μg/ml of penicillin-streptomycin (Wisent). 293T-ACE2 cell line was previously reported [24].

### Enzyme-linked immunosorbent assay (ELISA)

All samples were tested for anti-N total immunoglobulin levels using an in-house anti-N ELISA. The assay protocol is similar to the anti-SARS-CoV-2 RBD ELISA previously developed by our group [25], except that recombinant N (Centre National en Électrochimie et en Technologies Environnementales Inc., Shawinigan, Canada) was used (0.25 µg/ml) in lieu of the RBD antigen (2.5 µg/ml).

### Cell surface staining and flow cytometry analysis

293T were transfected with full-length SARS-CoV-2 Spikes and a green fluorescent protein (GFP) expressor (pIRES2-eGFP) using the calcium-phosphate method. Two days post-transfection, Spike-expressing 293T cells were stained with the CV3-25 Ab (5 μg/mL) as control or plasma (1:250 dilution) for 45 min at 37°C. AlexaFluor-647-conjugated goat anti-human IgM+IgG+IgA (1/800 dilution) were used as secondary Abs. The percentage of Spike-expressing cells (GFP + cells) was determined by gating the living cell population based on viability dye staining (Aqua Vivid, Invitrogen). Samples were acquired on a LSRFortessa cytometer (BD Biosciences), and data analysis was performed using FlowJo v10.7.1 (Tree Star). The conformationally-independent anti-S2 antibody CV3-25, effective against all Spike variants, was used to normalize Spike expression, as reported [4,20,22,23]. The Median Fluorescence intensities (MFI) obtained with plasma were normalized to the MFI obtained with CV3-25 and presented as percentage of CV3-25 binding.

### Virus neutralization assay

293T cells were transfected with the lentiviral vector pNL4.3 R-E− Luc and a plasmid encoding the D614G or the BQ.1.1 S glycoprotein at a ratio of 10:1 to produce SARS-CoV-2 pseudoviruses. Two days post-transfection, cell supernatants were harvested and stored at −80°C until use. For the neutralization assay, 293T-ACE2 target cells were seeded at a density of 1×10^4^ cells/well in 96-well luminometer-compatible tissue culture plates (PerkinElmer) 24h before infection. Pseudoviral particles were incubated with several plasma dilutions (1/50; 1/250; 1/1250; 1/6250; 1/31250) for 1h at 37°C and were then added to the target cells followed by incubation for 48h at 37°C. Cells were lysed by the addition of 30 μL of passive lysis buffer (Promega) followed by one freeze-thaw cycle. An LB942 TriStar luminometer (Berthold Technologies) was used to measure the luciferase activity of each well after the addition of 100 μL of luciferin buffer (15mM MgSO_4_, 15mM KH_2_PO_4_ [pH 7.8], 1mM ATP, and 1mM dithiothreitol) and 50 μL of 1mM d-luciferin potassium salt (Prolume). The neutralization half-maximal inhibitory dilution (ID_50_) represents the plasma dilution to inhibit 50% of the infection of 293T-ACE2 cells by pseudoviruses.

### Statistical analysis

Symbols represent biologically independent samples from individuals. Statistics were analyzed using GraphPad Prism version 8.0.1 (GraphPad, San Diego, CA). Each dataset was tested for statistical normality and this information was used to apply the appropriate (parametric or nonparametric) statistical test. p values < 0.05 were considered significant; significance values are indicated as ^*^P<0.05, ^**^P<0.01, ^***^P<0.001, ^***^P<0.0001, ns, non-significant.

## Results

We first monitored the capacity of plasma to recognize the D614G and BQ.1.1 Spikes after the third and fourth doses of mRNA vaccine by flow cytometry (Figure 1B-D). For the D614G S, no significant differences were observed four weeks and four months after the third dose of vaccine between individuals with or without recent BTI. In contrast, four weeks after the fourth dose of mRNA vaccine, individuals with recent BTI recognized better the D614G S than donors with no recent BTI regardless of the vaccine type received (Figure 1B). For the BQ.1.1 S, at the M4-Va3 timepoint donors with recent BTI better recognized the S than individuals with no recent infection, and this difference in recognition was more significant four weeks after the fourth dose (Figure 1C). The level of BQ.1.1 S recognition was significantly lower compared to D614G S in donors without recent BTI (Figure 1D), in agreement with recent reports [4,5]. In donors who had recently been infected, there was a significant but smaller difference in the level of recognition between the two Spikes compared to the other group.

We also measured the neutralizing activity of plasma against the D614G and BQ.1.1 S (Figure 1E-G). We observed pattern similar to that measured for Spike recognition. No significant differences were observed between the two groups at W4-Va3 and M4-Va3 timepoints (Figure 1E-F). In contrast, four weeks after the fourth dose, donors with recent BTI had a significantly higher level of neutralizing activity against D614G and BQ.1.1 S. All donors with recent BTI who received a fourth dose developed neutralizing antibodies against BQ.1.1 S, while some donors who just received four doses of vaccine were still not able to neutralize this Spike. As observed for S recognition (Figure 1D), BQ.1.1 Spike was significantly less neutralized than D614G S, even after four doses of mRNA vaccine (Figure 1G). However, the difference in neutralization between the two S was smaller in the group with recent BTI.

The Moderna BA.1 bivalent vaccine (blue points) tended to induce better recognition and neutralization than the other vaccine platforms including the Pfizer BA.4/5 bivalent vaccine with lesser decrease of recognition and neutralization of BQ.1.1 S (Figure 1B-G, Figure S2). These differences did not reach statistical significance; whether this is due to the relatively low number of samples tested remains to be determined.

## Discussion

Since its emergence in late 2021, the Omicron variant continues to evolve into new subvariants that are increasingly resistant to monoclonal antibodies and vaccination [5,26–30]. To address vaccine resistance, bivalent mRNA vaccines, expressing both the original Spike and one of the parental lineages of Omicron (BA.1 or BA.4/5) Spike, have been developed and are now being administered in several jurisdictions worldwide. However, although the bivalent mRNA vaccine has been shown to increase the level of protection against BA.5 variant in mice [31], evidence of its superior effectiveness in the human population remains to be demonstrated, especially against sub-lineages with newly acquired immune escape mutations. Recent studies showed that both monovalent and bivalent vaccines induced low humoral responses against BQ.1.1, but recent breakthrough infection before vaccination strongly improved these responses [32]. The results presented herein support these observations.

As previously reported in numerous studies, including ours, hybrid immunity led to better humoral responses against the BQ.1.1 and other recent variants than just vaccination [4,5,32]. Also, we observed that after 4 doses of mRNA vaccine and no recent BTI, some donors did not have neutralizing activity against pseudoviral particles bearing the BQ.1.1 Spike. Whether these changes of recognition and neutralization translate into greater risk of severe disease is currently unknown. In contrast, BTI likely increased the breadth of neutralizing antibodies since all donors had detectable levels of neutralization against BQ.1.1.

These results indicate that further efforts have to be devoted to improve vaccines against new SARS-CoV-2 variants of concern. Whether immune responses comparable to those observed with breakthrough infections could be obtained with new vaccine formulations remains to be determined.

## Data Availability

All data produced in the present study are available upon reasonable request to the authors

## Author Contributions

A.T., R.B. and A.F. conceived the study. A.T., M.B., H.M., J.P., G.G-L., R.B. and A.F. performed, analyzed, and interpreted the experiments. A.T. performed statistical analysis. H.M., G.G-L., M.C., and A.F. contributed unique reagents. L.G., P.A., C.M., C.T., D.E.K., Y.G. and V.M.-L. collected and provided clinical samples. R.B., G.D.S., and I.L. provided scientific input related to VOCs and vaccine efficacy. A.T. and A.F. wrote the manuscript with inputs from others. Every author has read, edited and approved the final manuscript.

## Informed Consent Statement

Informed consent was obtained from all subjects involved in the study.

## Data Availability Statement

Further information, data reported in this paper, and requests for resources and reagents should be directed to and will be fulfilled by the lead contact, Andrés Finzi upon request.

## Acknowledgments

The authors are grateful to the donors who participated in this study. The authors thank the CRCHUM BSL3 and Flow Cytometry Platforms for technical assistance.

---

**Figure S1.**
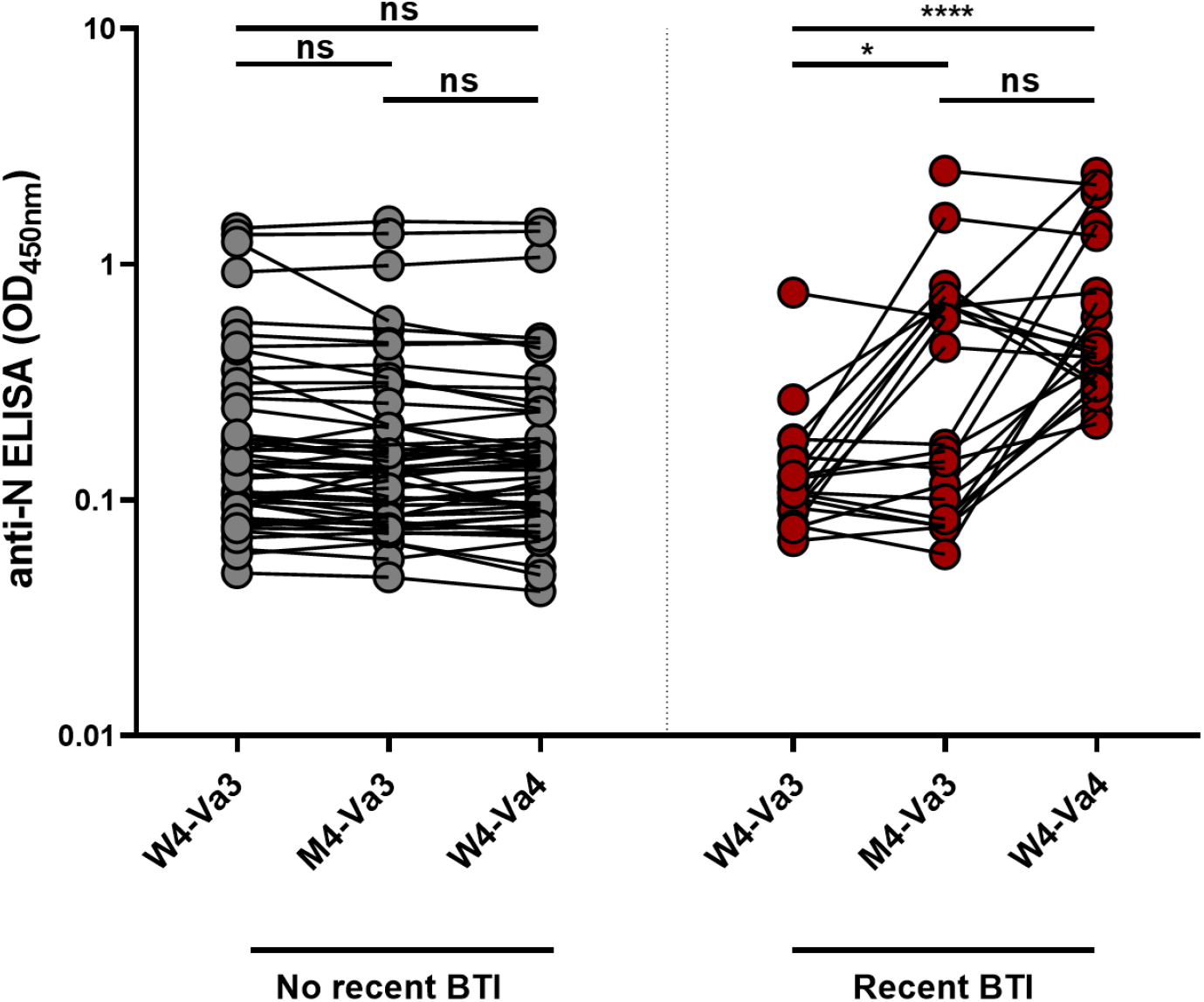
Anti-N level measured after the third and fourth doses of SARS-CoV-2 vaccine. Anti-N level was measured in plasma from vaccinated donors by ELISA. Donors are considered to have a recent BTI when a significant increase of anti-N Abs level between W4-Va3 and M4-Va3 or between M4-Va3 and W4-Va4 is observed, according to a recently described analytical approach based on the ratio of anti-N absorbance. Individuals with no recent BTI and recent BTI are represented by gray and red dots respectively. (* P < 0.05; **** P < 0.0001; ns, non-significant).

**Figure S2.**
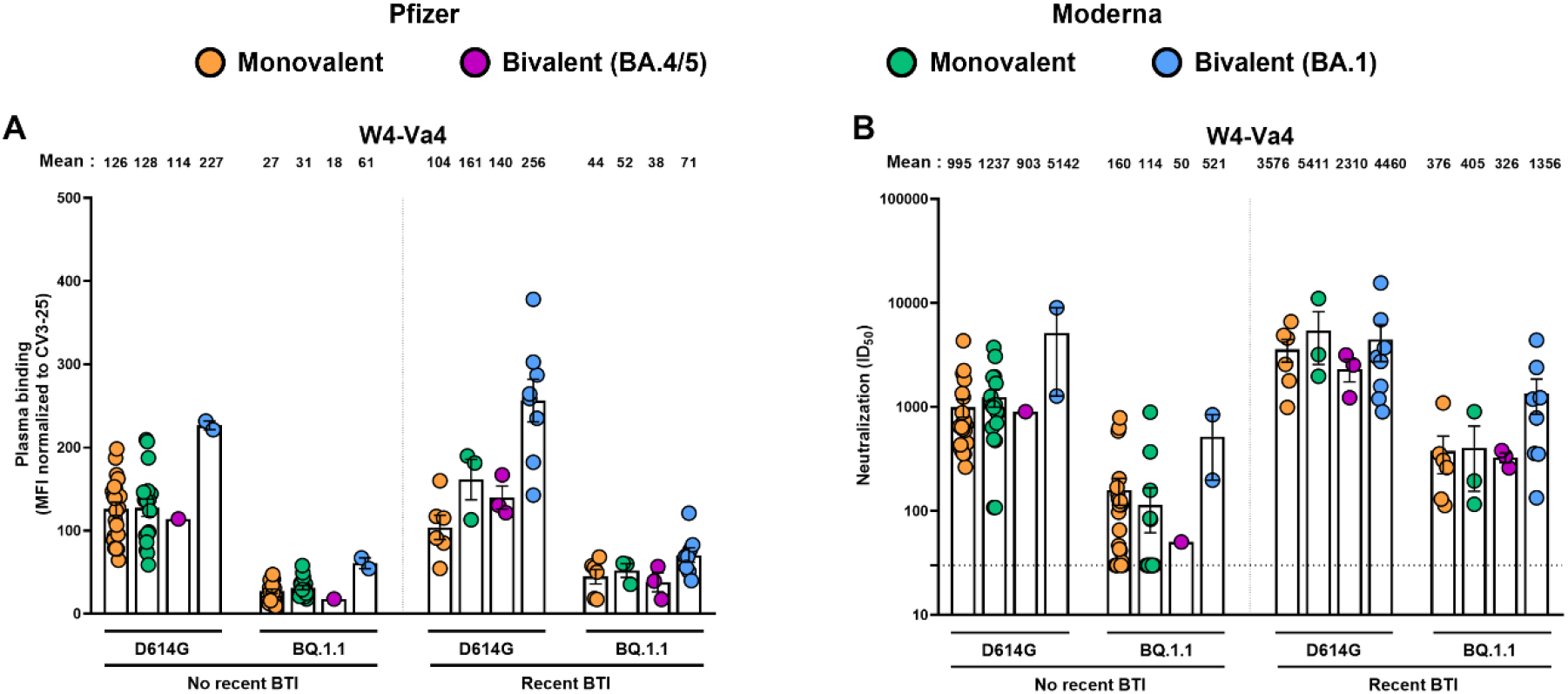
Recognition and neutralization of the D614G and BQ.1.1 Spikes after the fourth doses of SARS-CoV-2 vaccine in individuals with or without a recent breakthrough infection. (**A**) 293T cells were transfected with the full-length D614G or BQ.1.1 S, stained with the CV3-25 mAb or with plasma from vaccinated individuals and analyzed by flow cytometry. The values represent the MFI normalized by CV3-25 mAb binding. (**B**) Neutralization activity was measured by incubating pseudoviruses bearing SARS-CoV-2 S glycoproteins, with serial dilutions of plasma for 1 h at 37°C before infecting 293T-ACE2 cells. Neutralization half maximal inhibitory serum dilution (ID_50_) values were determined using a normalized non-linear regression using GraphPad Prism software. Individuals vaccinated with Pfizer monovalent, Moderna monovalent, Pfizer bivalent (BA.4/5) or Moderna bivalent (BA.1) fourth dose are represented by orange, green, purple and blue points respectively. Limits of detection are plotted. Error bars indicate means ± SEM.

